# Athletes’ access to, attitudes towards, and experiences of help-seeking for mental health: A scoping review protocol

**DOI:** 10.1101/2022.02.18.22271182

**Authors:** Kirsty R. Brown, Mary L. Quinton, Jennifer Cumming

**Affiliations:** School of Sport, Exercise and Rehabilitation Sciences, University of Birmingham, UK; Institute of Mental Health, University of Birmingham, UK

**Author notes:** **Corresponding author:** Kirsty R. Brown. **<u>Author contributions:</u>** KB conceived the research question for the scoping review with supervision from MQ and JC who approved and refined the idea. KB conducted literature searches and drafted the entire manuscript with input from all co-authors. MQ and JC provided feedback on the manuscript. All authors reviewed and approved the manuscript. **<u>Funding stated:</u>** This research received no specific grant from any funding agency in the public, commercial or not-for-profit sectors. **<u>Competing interests:</u>** All authors have completed the ICMJE uniform disclosure form at www.icmje.org/coi_disclosure.pdf and declare: that the Article Processing Charges will be covered by the University of Birmingham if accepted for publication; no financial relationships with any organizations that might have an interest in the submitted work in the previous three years; no other relationships or activities that could appear to have influenced the submitted work.

**Keywords:** Sport, well-being, mental illness, help-seeking behavior, treatment-seeking, mental health service, formal support

## Abstract

**Introduction:** Athletes are not immune to mental health issues but are less likely to seek help than nonathletes and experience barriers including lack of access to services, lack of knowledge as to how to access services and negative past experiences for help-seeking. Formal and semi-formal sources of support (e.g. support provided in healthcare, sporting context, and higher education systems) are key places for athletes to seek help for mental health, and there is a need to synthesise the evidence on athletes’ access, attitudes to, and experiences of, these services. This protocol outlines a scoping review that will be used to map the evidence, identify gaps in the literature, and summarise findings on athletes’ access, attitudes to and experiences of help-seeking for their mental health.

**Methods and analysis:** The methodological frameworks of Arksey and O’Malley (2005), Levac et al. (2010), and the Joanna Briggs Institute (2020 & 2021) were used to inform this scoping review protocol alongside the PRISMA-P checklist and published scoping review protocols within sport and health. This protocol outlines the background evidence, need for this scoping review, and the steps that will be taken.

**Dissemination:** The evidence will be mapped numerically and thematically to describe studies and highlight key concepts, themes, and gaps in the literature. The published scoping review will be disseminated to relevant stakeholders and policymakers including those in healthcare, the sporting context, and the higher education system. The resulting outputs will be in the form of both peer-reviewed and non-peer reviewed publications (e.g. multimedia in the form of a blog post and at conferences).

**Strengths and limitations:**

- Strength: The protocol outlines a novel scoping review that will contribute to a gap in the literature, and impact further research directions.
- Strength: Informed by best practice methodological frameworks to ensure rigour.
- Limitation: Lack of quality assessment of papers.
- Limitation: The review will only focus on formal and semi-formal sources of help-seeking (e.g. healthcare, the sporting context and higher education) and will not include research on informal or self-help sources.

## Introduction

In contrast to the commonly held perception that athletes are mentally strong, resilient, and do not experience mental ill health, it is increasingly evident that athletes are not immune to mental health issues. In samples of French and Australian athletes, prevalence rates were 17% and 46% respectively, with the most common disorders including anxiety, depression and eating disorders.[1,2] Despite these rates being similar or higher to those found in the general population, athletes have comparatively much lower rates of help-seeking for mental health.[3,4]

The mismatch between prevalence of mental health issues and rates of help-seeking in athletes is of concern. Help-seeking is “the process of actively seeking out and utilising social relationships, either formal or informal, to help with personal problems.”[5](p.8) In a study of Norwegian elite athletes, 13.5% presented with an eating disorder compared to 4.6% in the general population.[6] While the rates of help-seeking for anorexia nervosa, bulimia and binge eating are found to be 34.5-62.6% in the general population,[3] the rate of help-seeking for eating disorders in athletes was 1.5%.[4] Thus, a significant proportion of athletes are likely not getting the help required. A scoping review is needed to map the literature on athletes’ attitudes towards and experiences of help-seeking, including how they access and utilise different forms of mental health services, and identify where evidence gaps exist.

There is a lack of conceptual and theoretical frameworks within the help-seeking literature.[7] However, Rickwood et al[5] and Rickwood and Thomas[7] have proposed two complimentary help-seeking frameworks which have informed the background to this scoping review, and the inclusion/exclusion criteria as will be discussed. The first framework suggests a 4-step process: 1) “awareness and appraisal of problems”, 2) “expression of symptoms and need for support”, 3) “availability of sources of help”, and 4) “willingness to seek out and disclose to sources.”[5](p. 8) The second framework proposed 5 main components: 1) process (“the part of the behavioural process that is of interest”), 2) timeframe (when the action occurs), 3) source (where the assistance for help is sought from), 4) type (“the form of actual support that is sought”), and 5) concern (“the type of mental health problem for which help is being sought”).[7](p. 180-182) Further details on these frameworks are provided in the appendix. For the purposes of this scoping review protocol, the background will be mapped onto step 3 (availability of sources of help) and 4 (willingness to seek out and disclose to sources), and the process and source components of Rickwood and colleagues frameworks.[5,7]

### Availability of sources of help (step 3): athletes and coaches perceived access to services[5]

As noted above, the third step in Rickwood et al’s framework is availability of sources of help.[5] A barrier to young people and students seeking help for their mental health includes a lack of physical access to services and knowledge of how to access services.[8–10] Similar results have been found in athlete populations.[11,12] In a sample of Canadian athletes, 47.3% chose not to seek services for mental health when desired, with lack of available services as the main reason.[11] In contrast, 98% of US collegiate sport coaches in Sudano and Miles’ (2017) study stated that student-athletes can access mental health care services.[13] Therefore, views on access to mental health services may differ between coaches and athletes as well as between contexts, highlighting the need for a scoping review to map and better understand athletes’ access to services.

### Willingness to seek out and disclose to sources (step 4): past experiences[5]

Step four in Rickwood and et al’s framework is willingness to seek out and disclose to sources.[5] Another key barrier for athletes seeking help for mental health is negative past experiences whereas a comparative facilitator is positive past experiences[12,14] This suggests that athletes do have experiences of mental health services, but there is a need to review the literature on athletes’ experiences of mental health help-seeking beyond the facilitators and barriers.

### General orientation or attitude toward obtaining assistance, and observable behaviour (process): preferences for help seeking[7]

When athletes do seek help, they are more likely to go outside the sport environment, and least likely to seek help from coaches.[11,15] This maps onto the process component in Rickwood and Thomas’ (2012) framework;[7] that is, where athletes seek help from can be understood as an observable behaviour. Additionally, their preferences for help-seeking can be understood as a general orientation or attitude toward obtaining assistance. Existing literature has found that athletes express preferences for sport psychologists, counselors, physiotherapists, and clinicians and place importance on these professionals understanding sport demands or just having the ability to value the role of sport in the athletes’ life.[16– 20] It is now well established that the therapeutic relationship/alliance between a patient and service provider impacts treatment outcomes.[21–24] It is important to consider athletes’ preferences for who to seek help from to ensure an optimal therapeutic alliance, and therefore treatment outcomes are achieved. However, there is yet to be a review that maps evidence on athlete preferences for a provider and their experiences of interacting with them, which is important to understand.

### Formal and semi-formal sources of help (source): e.g. healthcare, the sporting context and the higher education system[7]

Aligning with Rickwood and Thomas’ third component (source), help-seeking within primary care is a formal source of help due to the “specified role in delivery of mental health care.”[7](p.181) Within UK primary care, sport psychiatry is not yet widely available.[25] Similarly, in Canada there is only one centre for providing athlete-specific mental health services: The Canadian Centre for Mental Health in Sport (CCMHS).[26,27] Therefore, athletes are likely utilizing mental health services provided for the general population.

Institutions and clinicians have been identified as key areas to address and improve athlete mental health.[28] However, it is still unclear which types of formal and semi-formal sources of support are most utilised by athletes struggling with their mental health. Within higher education, for example, both formal (e.g., counsellors) and semi-formal sources (e.g., academic tutors) of help are available. Similarly, there are both formal (e.g. GP, psychologist and psychiatrist) and semi-formal (e.g. physiotherapist and dietician) sources of support in healthcare. In the sporting context, a coach and manager can be understood as semi-formal sources of support. It is important to map the existing literature on athlete interactions and experiences with formal and semi-formal sources of support for their mental health such as those within healthcare, the sporting context and the higher education system.

### Aims of this scoping review

In sum, there is a need to synthesise the literature on athletes’ mental health help-seeking in the form of a scoping review to improve their help-seeking experiences and mental health outcomes. This paper outlines the protocol for a scoping review that will aim to assess and map: 1) The literature on athlete access, attitudes to and utilization of formal sources of mental health support, 2) The literature on athlete experiences of mental health help-seeking from formal sources of support, 3) Current gaps in the literature, and 4) What the literature recommends as further research. A scoping review is appropriate for addressing these aims because athlete mental health within sport psychology is a relatively new yet growing area of research, with evidence continually emerging.[29]

## Methods and analysis

### Frameworks to inform the scoping review

As is common practice in scoping reviews, a number of methodological frameworks and recommendations have been used to inform this protocol, alongside published scoping review protocols within sport and health.[29–38] The 5 stage framework proposed by Arksey and O’Malley (2005) was the predominant framework utilised.[31] This was enhanced by Levac and colleagues (2010) who have provided further recommendations based on each stage of this framework.[29] Additionally, the Joanna Briggs Institute (JBI) (2020 and 2021) framework and recommendations were used to ensure that this scoping review meets their stated purpose as well as providing the Person-Concept-Context (PPI) to inform the title.[32,33] To ensure rigor the PRISMA-P checklist also informed this protocol and will inform the scoping review.[34]

Rickwood and Thomas’s (2012) framework informed the inclusion and exclusion criteria.[7] Both of Rickwood and colleagues’ frameworks will aid the data analysis and discussion.[5,7]

### Stage 1: Identifying the research question

To identify the research question (i.e., Athletes’ access to, attitudes towards and experiences of help-seeking for mental health) and the inclusion and exclusion criteria, JBI’s PCC (Person-Concept-Context) framework was applied alongside the help-seeking frameworks.[5,7,32] Table 1 shows how these frameworks were used to inform the research question. Further details on the two conceptual frameworks provided by Rickwood and colleagues, and how it maps onto the research question are provided in the appendix (see appendix 1: tables 1 and 2).

**Table 1:** Research question and how it maps onto conceptual frameworks[5,7,32]

| Component of research question | PPC framework | Rickwood et al (2005) | Rickwood & Thomas (2012) |
| --- | --- | --- | --- |
| Athletes | Person |  |  |
| access to | Concept | Availability of sources of help |  |
| attitude towards | Concept | Willingness to disclose to sources |  |
| and experiences of | Concept | Willingness to disclose to sources |  |
| Help-seeking for mental health | Concept | Willing to seek out sources | Type: emotional support, treatment/health service provision, & informational support<br>Concern: any mental health concern; specific symptoms and general distress<br>Source: formal and semi-formal |

### Stage 2: identifying relevant studies

As recommended, the inclusion and exclusion criteria have been initially predetermined.[29,31] As the scoping review is carried out, and the literature is assessed, the criteria will be reassessed and modified as required. The decisions on inclusion and exclusion criteria will involve a research team,[29] consisting of 3 individuals (KB, MQ, and JC) with experience in conducting systematic reviews.

Primarily the inclusion criteria will be:

- Athletes from any sport (person)[32]
- Athletes aged 16+ (person)[32]
- All genders of athletes (person)[32]
- Process:
  - Observable mental health help-seeking behaviour (in the past). For example, going to see a primary care clinician for mental health concerns
  - General orientation or attitude toward obtaining assistance. For example, preference for the source of help for a mental health concern
  - Future behavioural intention to gaining support. For example, where the athlete with gain support for a mental health concern in the future[7]
- Timeframe: ever (e.g. past behaviours and future intentions)[7]
- Source: formal and semi-formal sources of mental health support[7]
  - E.g. university counsellors and welfare officers in higher education, and a general practitioner, psychologist and psychiatrist in healthcare (formal)
  - E.g. university lecturer, academic tutor and sports coach in higher education, and physiotherapist and dietician in healthcare (semi-formal)
  - E.g. sports coach and manager in the sporting context (semi-formal)
- Type: emotional support, treatment/health service provision, and informational support[7]
- Concern: any mental health concern: specific symptoms & general distress[7]
- Primary research, reviews of any type (e.g. systematic, scoping) (this is a scoping review of the published literature)
- Papers in English

The exclusion criteria will be as follows:

- Source: semi-formal, informal, or self-help[7]
- Type: instrumental support and affiliative support[7]
- Opinion pieces, magazine articles, grey literature, and newspaper articles
- Papers in languages other than English

#### Stage 2a: Initial preliminary searches

Initial searches will be carried out using relevant terms including: athlete, help-seeking experiences, access to mental health services, support in education, and mental health. Relevant search strategies from similar systematic and scoping reviews will be used.(e.g.[12,39,40]) Following an assessment of literature, and a discussion between the authors, other colleagues within the research group and with a research librarian, key words will be identified.

#### Stage 2b: identification of key words and terms

Following the initial searches, discussions and review of the papers, a detailed search strategy will be produced. The search strategy will use the Boolean operators of AND and OR. Truncations will also be applied where appropriate. A draft search strategy for the OVID platform is provided in the appendix (see appendix 2).

Once the search strategy has been developed the databases to be searched will be: PsychINFO (via OVID), Embase (via Ovid), MEDLINE (via Ovid and Web of Science), APA PsychArticles Full Text (via OVID), Web of Science core collection, Sport Discus (via EBSCO), CINAHL (via EBSCO), Proquest (education database), Proquest (health and medical collection), Proquest (Nursing and Allied Health database), Proquest (psychology database), Proquest (public health database), and Proquest (Sports Medicine & Education). These databases will be modified as needed during the review process.

#### Stage 2c: searching of references and citations

Again, the reference lists of identified studies will be searched until the point when no more new studies are being retrieved.[31]

### Stage 3: Study selection

#### Stage 3a: title and abstract screening

Firstly, the titles and abstracts of all identified studies will be reviewed. Similarly to the protocol published by Griffin and colleagues,[35] the lead author/reviewer (KB) will review all titles and abstracts. The second reviewer (MQ) will review a random selection of 30% of the titles and abstracts. At a minimum, the reviewers meet at the beginning, midpoint and end of the abstract review[29,31,32] to discuss the eligibility of studies where a conflict may have arisen, and any changes to the inclusion criteria they would like to make. If disagreements occur on the eligibility of a study or changes the inclusion criteria, then the third reviewer (JC) will be involved.

If full texts are not available, then authors will be contacted to obtain the article.

#### Stage 3b: review of full articles for inclusion

Following title and abstract screening, the full articles will then be reviewed. The lead reviewer (KB) will review all of the full texts, and the second reviewer (MQ) will review 20%. Again, authors will meet frequently and the third reviewer (JC) will be involved to resolve disagreements, should they occur.

Should an agreement not be reached, then the article will be excluded from the review.

If further information from included studies is required, then authors will be contacted to obtain supplementary material or clarification.

### Stage 4: Charting the data

To collect the relevant data, a data charting form will be created in Covidence.[31,41] This form will be updated as the research process and data extraction occurs, through discussion between the reviewers.[29]

#### Stage 4a: testing the data charting form

In line with recommendations by Levac and colleagues,[29] this data charting form will be tested by two reviewers (KB and JC). They will independently extract data from 5 studies and put it onto the data charting form in Covidence. All reviewers (KB, JC and MQ) will then meet to discuss the data charting form, and any changes required.

#### Stage 4b: charting the data from all included studies

Once the initial data charting form has been agreed upon, the data extraction process will commence. Data will be extracted and charted from all included studies by the lead author (KB) and then 20% will be independently extracted by the second reviewer (JC). Disagreements in the charting of data will be discussed by all three reviewers (KB, JC, and MQ) to resolve conflicts.[35]

Based on other scoping reviews and the frameworks used to inform our scoping review,[29,31–33,35– 38] we will likely extract the following information onto our data charting form. An example of the data extraction form as it appears on Covidence is displayed in the appendix (see appendix 3).

1. Title
2. Authors (including corresponding author details)
3. Year of publication
4. Location in which the study was conducted (country, city, institution)
5. Type of study (systematic review, scoping review, primary article or grey literature)
6. Size of study population
7. Study population (e.g. age, gender, sport, level of competition)
8. Inclusion and exclusion criteria
9. Gap in the literature that the study fills
10. Aims of the study
11. Method of recruitment of participants (e.g. phone, email, poster)
12. Data collection method (quantitative, qualitative or mixed methods)
13. Details of data collection (e.g. questionnaires used for quantitative)
14. Intervention (if appropriate)
15. Details of intervention (e.g. duration of intervention, outcome)
16. Key findings that relate to the aims of the scoping review. (e.g. is the focus on access/attitudes/experiences)
17. Gaps identified (e.g. source of support sought)
18. Future directions identified (e.g. recommendations for a systematic review)

### Stage 5: collating, summarising and reporting the results (data analysis plan)

The data analysis plan is to collate, summarise and report the results in two main steps: 1) Descriptive numerical description, and 2) Thematic analysis.

#### Stage 5a: descriptive numerical

This is a descriptive and numerical analysis of all studies included.[29,31,32] This description will include steps such as: the overall number of studies, the type of the study, the year of publications, characteristics of study population and the number of studies from particular countries. For qualitative studies, the main results and gaps identified will be reported.

#### Stage 5b: thematic analysis

Thematic analysis is recommended by Arksey and colleagues and the Joanna Briggs Institute frameworks.[31,32] This will include the identification of themes in the studies.

#### Stage 5c: presentation of the data

As recommended by the Joanna Briggs Institute we will firstly describe the search strategy and selection process, and provide a PRISMA diagram.[32]

#### Descriptive numerical

We will present this data in tables,[32] separated by appropriate topic within the scoping review. For example, 1) a table for athlete access to formal and semi-formal sources of support for their mental health, and 2) a table for athlete attitudes towards formal and semi-formal sources of support for their mental health. A final table will present overall descriptive statistics (e.g. frequency counts of concepts and frequency of country studies).

#### Qualitative

The qualitative studies will be presented in thematic analysis tables which will be grouped by themes, gaps and further research directions. An example of these thematic analysis tables is displayed in the appendix.

The two conceptual frameworks by Rickwood et al (2012) and Rickwood and Thomas (2005) will aid the presentation of the data, themes, and the overall discussion within the scoping review.[5,7]

### Optional stage 6: Consultation exercise (public and patient involvement)

Arksey and O’Malley suggest an optional stage of consultation and stakeholder involvement.[29,31] Levac and colleagues stress the importance of this step and provide further recommendations on how to best involve stakeholders.[29] We agree with the importance of public and patient involvement in health research, but unfortunately time will not allow it in the case of our scoping review. However, public and patient involvement (PPI) will be used to inform later research on this topic.

## Supporting information

PRISMA Scoping review checklist

## Data Availability

All data produced in the present work are contained in the manuscript

## Ethics and dissemination

The findings of this scoping review will be presented to the Patient and Public Involvement (PPI) group involved with this research. The results will then be published in peer-reviewed journals, presented at conferences and summarised in non-academic formats. For example, in the form of a blog.

## Acknowledgements

We would like to thank Lynne Harris from the University of Birmingham library services who provided feedback on the search strategy, and selection of databases.

## Notes

### Funding Statement

This study did not receive any funding.
Article Processing Charges will be covered by the University of Birmingham if accepted for publication

