## Supplemental Appendix for "Athletes’ access to, attitudes towards, and experiences of help-seeking for mental health: A scoping review protocol"

**Appendix 1) Conceptual frameworks:**

**Rickwood et al (2005)**

| Step of framework | Is this a focus of the scoping review? | How it maps onto the scoping review & inclusion criteria |
| --- | --- | --- |
| 1) Awareness and appraisal of problems | No |  |
| 2) Expression of symptoms and need for support | No |  |
| 3) Availability of sources of help | Yes | **Access** to mental health support from formal and semi-formal sources *e.g. within healthcare, the sporting context and the higher education system* |
| 4) Willingness to seek out and disclose to sources | Yes | **Attitudes** towards help for mental health from formal and semi-formal sources *e.g. within healthcare, the sporting context and the higher education system* |

*Table 1: Rickwood and et al (2005) help-seeking framework and how it maps onto this scoping review and the inclusion criteria*

**Rickwood and Thomas (2012)**

| Component of framework & definition of component | Details of framework component | Is this a focus of the scoping review? | How it maps onto the scoping review & inclusion criteria |
| --- | --- | --- | --- |
| Process: "Refers to the part of the behavioural process that is of interest."(p.180) | Orientation: "general orientation or attitude towards obtaining assistance"(p.180) | Yes | Athletes’ attitudes towards formal and semi-formal sources of mental health support    Athlete preferences for help-seeking |
|  | Intention: "future behavioural intention"(p.180) | No |  |
|  | Behaviour: "observable behaviour either in the past or prospectively in the future"(p.180) | Yes | Which formal and semi-formal sources of support athletes have sought help for their mental health |
| Timeframe: "A course of action takes place within a particular time frame, and this needs to be specified clearly. The better defined the time frame, the better respondents are able to provide a reliable and valid response."(p.181) | I.e. past/next 4 weeks | No |  |
|  | Past/next 12 months | No |  |
|  | Ever | Yes | Any past help-seeking behaviour from formal and semi-formal sources of support such as those in healthcare, the sporting context and higher education system |
| Source: "Source refers to the source of the assistance that is sought. Sources vary according to the level of professional expertise of the source and the relationship with the person seeking help, as well as the medium of the source (eg, online)."(p.181) | Formal: "professional health service providers with a specified role in delivery of mental health care (formal), ie, psychiatrist, psychologist, general practitioner, mental health nurse"(p.181) | Yes | For example, formal sources of support within healthcare: general practitioner, psychologist and psychiatrist  For example, formal sources of support within higher education: university counsellors and welfare officers |
|  | Semi-formal: "service providers and professionals who do not have a specified role in delivery of mental health care (semiformal), ie, teacher, work supervisor, academic advisor, youth worker, coach"(p.181) | Yes | For example, semi-formal sources of support within healthcare: physiotherapist and dietician    For example, semi-formal sources of support within higher education: university lecturer, academic tutor and university sports coach  For example, semi-formal sources of support within the sporting context: coach and manager |
|  | Informal: "informal social supports (informal), ie, friend, partner, parent"(p.181) | No |  |
|  | Self-help: "self-help resources (self-help), ie, unguided website use."(p.181) | No |  |
| Type: "Type of assistance refers to the form of actual support that is sought, such as psychoeducation, referral, supportive counseling, and therapy"(p.181) | Instrumental support: “financial assistance, transport"(p.182) | No |  |
|  | Informational support: “health-related information, referral information"(p.182) | Yes | Mental health information given to athletes by sources of support within healthcare, the sporting context and higher education system    Referral information given to athletes by sources of support within healthcare, the sporting context and higher education system |
|  | Affiliative support: “I.e. peer support”(p.182) | No |  |
|  | Emotional support: “support for emotional wellbeing”(p.182) | Yes | Emotional support given to athletes by formal and semi-formal sources of support sources |
|  | Treatment: “A further category for the mental health context could be type of treatment or health service provision."(p.182) | Yes | Athlete access to mental health treatment from formal and semi-formal sources of support |
| Concern: “A further category for the mental health context could be type of treatment or health service provision."(p.182) | General distress/concern | Yes | The concern an athlete seeks mental health support for can be general and not a specific mental health symptom |
|  | Specific symptom types I.e. depression | Yes | The concern an athlete seeks mental health support for can be a specific mental health symptom |

*Table 2: Rickwood and Thomas (2012) conceptual help-seeking framework, if it is the focus of the scoping review, and how it maps onto this scoping review and the inclusion criteria*

**Appendix 2) Draft search strategy: OVID**

exp athlete/ or athlete.mp.

**AND**

exp well being/ or well being.mp.

OR

exp mental health/ or mental health.mp.

OR

exp Mental Disorders/ or Mental disorder.mp.

OR

exp Mental illness/ or Mental illness.mp.

**AND**

exp help seeking/ or help seeking.mp.

OR

exp treatment seeking/ or treatment seeking.mp.

OR

exp help/ or help.mp.

OR

exp Mental Health Service*/ or Mental health service*.mp.

**Appendix 3: COVIDENCE draft data extraction form:**

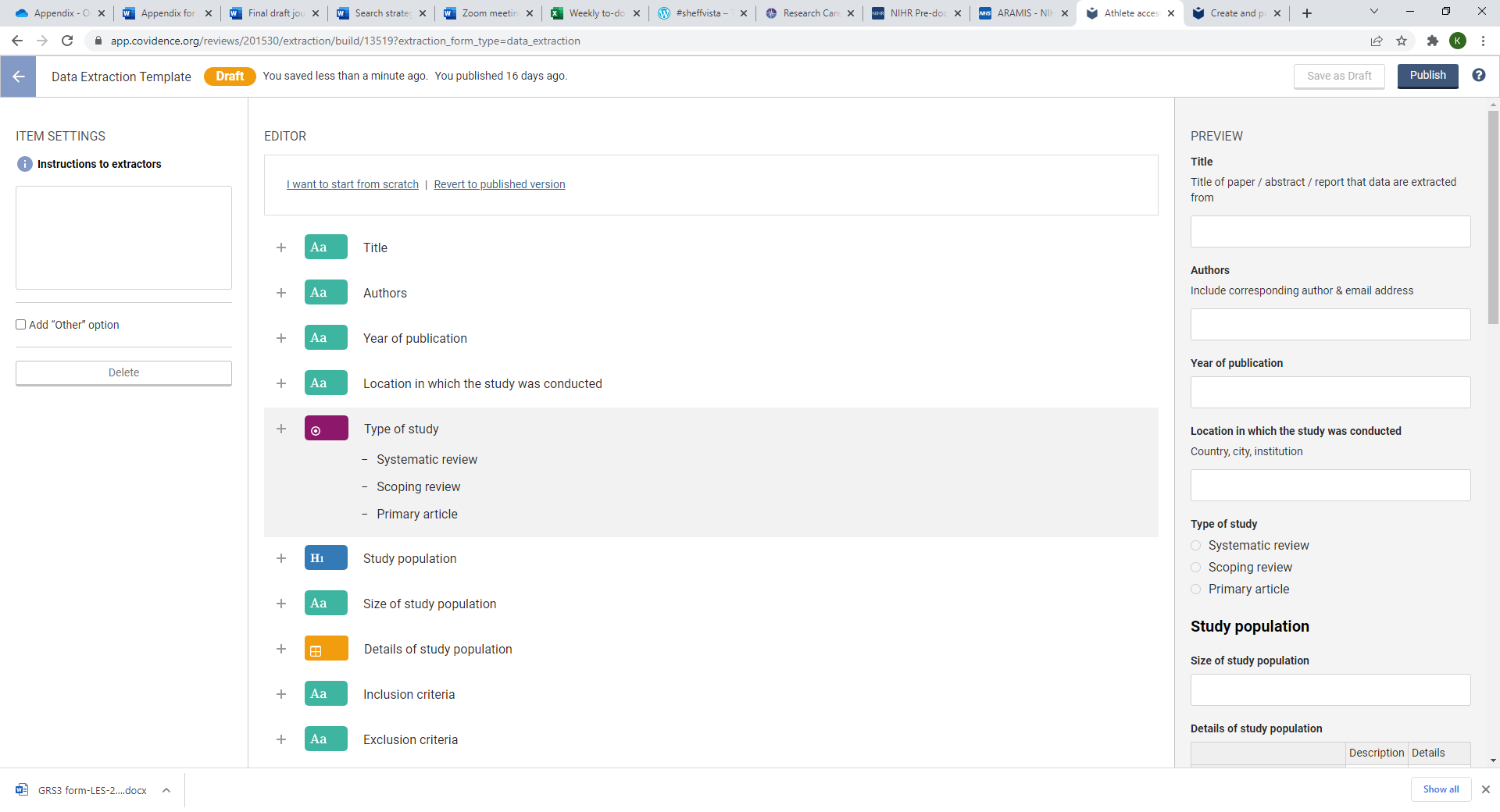

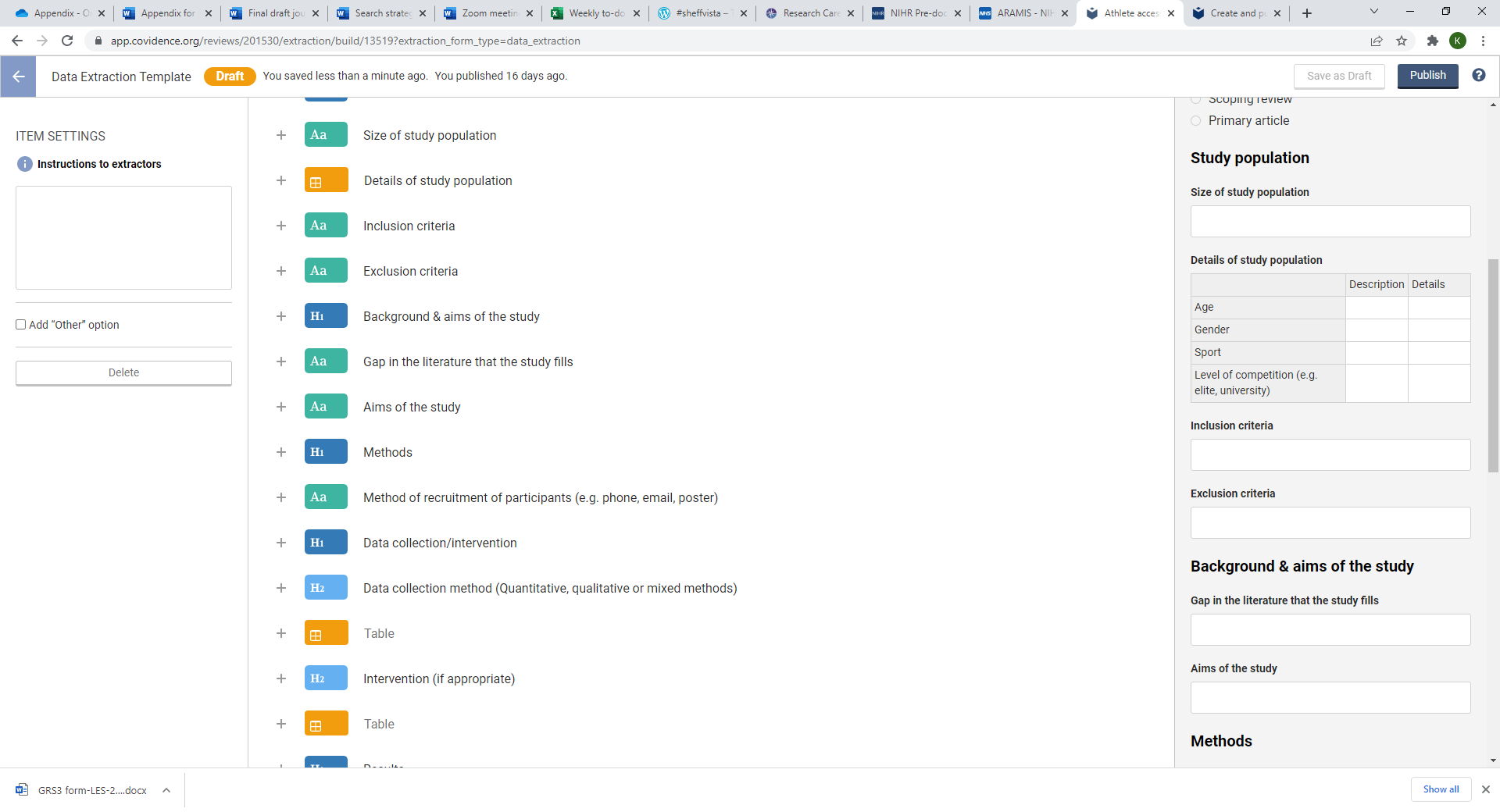

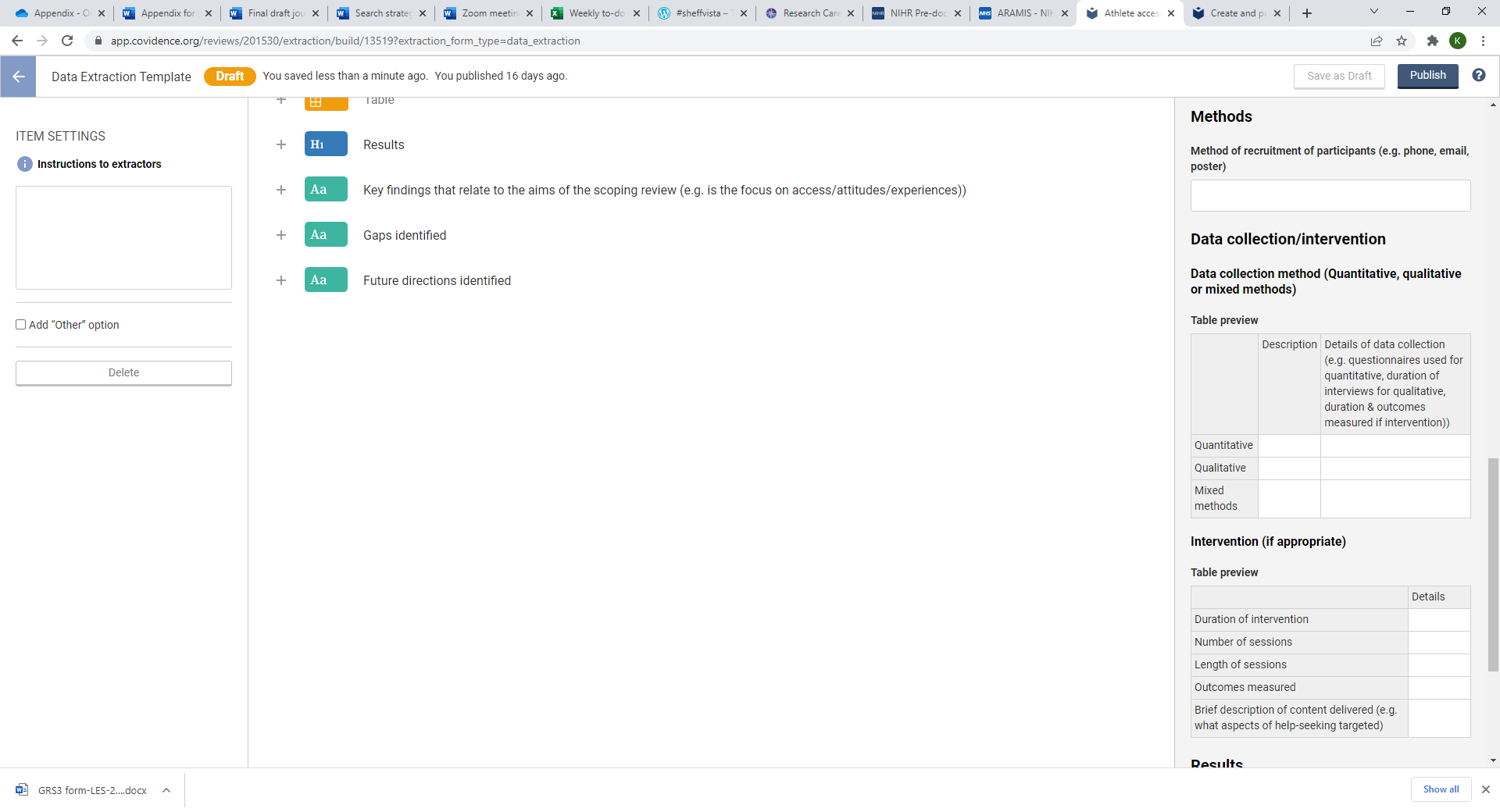

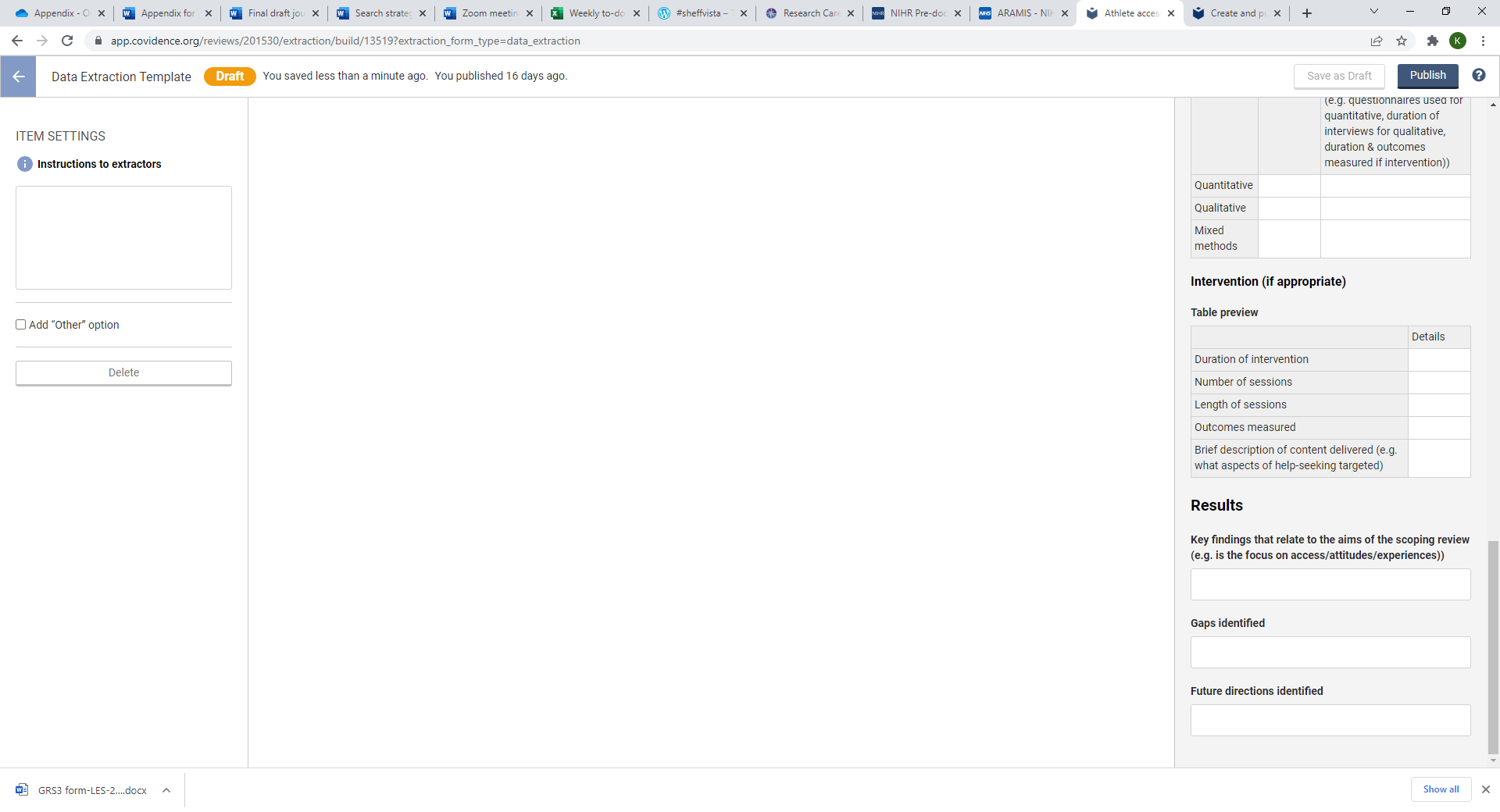
